# Wastewater Surveillance Reveals Testing-Related Underreporting and Hospital-Acquired SARS-CoV-2 Infections

**DOI:** 10.1101/2025.09.16.25335846

**Authors:** Michio Murakami, Nobuhisa Ishiguro, Hiroki Ando, Mutsumi Ishida, Toshihiro Hamada, Sho Nakakubo, Reiko Oyamada, Takahiro Hayashi, Yusuke Niinuma, Keisuke Kagami, Tatsuya Fukumoto, Keisuke Taki, Tomoyuki Endo, Masaaki Kitajima

## Abstract

Wastewater surveillance to monitor the incidence of infections faces challenges, in terms of discrepancies with sentinel-confirmed cases. We examined whether testing rates could explain the discrepancy between SARS-CoV-2 RNA found in a City of Sapporo wastewater treatment plant and the number of infections recorded at Hokkaido University Hospital over a period of approximately four years. Then, we analyzed the association between wastewater RNA concentrations with incidences of new cases among hospital-acquired infections. Linear regression analyses were performed using wastewater RNA concentrations as the independent variable and infected cases with and without correction for the testing rate as the dependent variable. In addition, modified Poisson regression analyses were performed, with the incidence of new cases among hospital-acquired infections as the dependent variable. After the legal reclassification of COVID-19 in Japan was changed to the same category as seasonal influenza, the rate of hospital testing declined significantly, though wastewater RNA concentrations remained high. Compared to non-correction for testing rates, corrected community-acquired infection cases showed a stronger association with wastewater RNA concentrations (*R^2^* = 0.54 and 0.75, respectively). The incidence of hospital-acquired infections was positively associated with wastewater RNA concentrations (incidence risk rate: 2.24 [95% confidence interval: 1.36–3.71]), and a log_10_ wastewater RNA concentration [copies/L] of 4.57 (4.10–5.03) was suggested as a 25% probability of new incidence. This study emphasized that SARS-CoV-2 wastewater surveillance is an objective and useful indicator reflecting infection incidence independent of testing rates.

## 1. Introduction

More than five years after the onset of the coronavirus disease 2019 (COVID-19) pandemic, individuals with underlying conditions such as cardiovascular disease, diabetes, chronic kidney disease, and malignancies, as well as older adults, remain at high risk of severe outcomes (Dryden-Peterson et al., 2025). With this in mind, it is important to monitor COVID-19 trends in communities, long-term care facilities, and hospitals.

Beginning February 1, 2020, COVID-19 was managed under Japan’s Infectious Diseases Control Law as a “designated infectious disease” (operationally, Category LJ–equivalent), and from February 13, 2021 onward, it was reclassified as a “pandemic influenza” infectious disease (also Category LJ–equivalent), a status that remained until May 7, 2023. On May 8, 2023, Japan reclassified COVID-19 as a Category LJ disease, aligning it with seasonal influenza. As a result, national case surveillance shifted from universal (all-case) reporting to sentinel surveillance (Sawakami et al., 2021). The costs of clinical testing and therapy have also become chargeable to patients.

Beginning in March 2020, several countries, including the Netherlands, the United States, Australia, and France, implemented wastewater surveillance for severe acute respiratory syndrome coronavirus 2 (SARS-CoV-2) to complement the limitations of sentinel surveillance, such as incomplete coverage of the population, potential reporting delays, and dependence on healthcare-seeking behaviors (Yang et al., 2023). In Japan, multiple municipalities have adopted a dual surveillance system of wastewater and sentinel surveillance since 2020, using wastewater data to complement sentinel data in assessing epidemic dynamics (Japan Institute for Health Security, 2025; Murakami et al., 2024).

In the City of Sapporo, Japan, wastewater surveillance for SARS-CoV-2 has been conducted since February 2021 (City of Sapporo, 2025). During the pandemic, SARS-CoV-2 RNA concentrations in wastewater closely tracked with confirmed cases in the city (Ando et al., 2023; Murakami et al., 2024) and at the hospitals (Kagami et al., 2023; Kagami et al., 2025). However, following the disease’s shift to endemic status, on May 8, 2023, a discrepancy between sentinel-confirmed cases and wastewater RNA levels progressed over time (Figure S1), as demonstrated previously (Kagami et al., 2025; Murakami et al., 2024). Such discrepancies have been documented in other settings that integrate wastewater surveillance with case-based sentinel surveillance, including in the United Kingdom, Denmark, Singapore, South Africa, Portugal, and Argentina (Iwu-Jaja et al., 2023; Jones et al., 2025; Krogsgaard et al., 2024; Masachessi et al., 2022; Monteiro et al., 2022; Wong et al., 2023).

Potential contributors to these discrepancies include: (1) increased proportions of asymptomatic or mild infections (Nakakubo et al., 2023), (2) reduced clinical testing (Boehm et al., 2023), (3) changes in healthcare-seeking behaviors (Maree et al., 2025), (4) variations in viral shedding patterns by age (Omori et al., 2021), and (5) differences in fecal excretion or environmental stability associated with emerging variants (Prasek et al., 2023; Sherchan et al., 2023). This study aimed to assess whether adjusting for the testing rate, corresponding to point (2) above, reduced clinical testing (Boehm et al., 2023), improves the association between wastewater RNA levels and confirmed cases during the endemic period. A previous study (Boehm et al., 2023) illustrated this issue by showing that, between late 2021 and early 2022, the widespread adoption of at-home antigen test kits, combined with the fact that positive results from these tests were not reported to public health authorities, led to a discrepancy between SARS-CoV-2 RNA concentrations in wastewater and the number of newly reported COVID-19 cases. The primary objective of this study was to investigate the discrepancy between wastewater surveillance and COVID-19 cases using a relatively small-scale but highly accurate dataset from a university hospital, comprising COVID-19 cases and the number of SARS-CoV-2 tests performed. We analyzed wastewater RNA concentration data from the City of Sapporo alongside COVID-19 cases and SARS-CoV-2 testing data from Hokkaido University Hospital. Furthermore, as a secondary objective, building on our previous finding of an association between wastewater SARS-CoV-2 RNA concentrations and the number of hospital-acquired COVID-19 cases among inpatients (Kagami et al., 2025), we examined whether wastewater SARS-CoV-2 RNA concentrations can be used to signal the incidence risk of hospital-acquired COVID-19 infections. A previous study (Kagami et al., 2025) reported weaker correlations between viral concentrations in wastewater with cases of hospital-acquired infections than with community-acquired cases. In this study, we focused on explaining the probability of new incidences using the viral concentrations in wastewater.

## 2. Materials and Methods

### 2.1. Ethical approval

Ethical approval for this study was obtained from the Institutional Review Board of Hokkaido University Hospital for Clinical Research (025-0012).

### 2.2. SARS-CoV-2 RNA analysis from wastewater samples

SARS-CoV-2 RNA concentrations in wastewater were obtained from a wastewater surveillance program conducted by the City of Sapporo. Other studies have also used data obtained from this program (Kagami et al., 2023; Kagami et al., 2025; Murakami et al., 2024). In this study, 24-hour composite untreated wastewater samples collected from Week 7 of 2021 (February 15–21, 2021) through Week 13 of 2025 (March 24–30, 2025), excluding Weeks 9–18 of 2023 and Week 1 of 2025, were used. Samples were collected from five catchment areas covered by three wastewater treatment plants in the City of Sapporo (each catchment area covers approximately 164,700 to 246,300 people, accounting for 52% of the city’s population of approximately 1.96 million), except from Weeks 7–14 of 2021, when samples were collected from three catchment areas. Sampling was generally conducted three times per week at each catchment area from Week 7 of 2021 to Week 39 of 2023, and once per week thereafter. Therefore, 9, 15, and 5 samples were collected per week from Weeks 7–14 of 2021, from Week 15 of 2021 through Week 39 of 2023, and from Week 40 of 2023 through Week 13 of 2025, respectively. SARS-CoV-2 RNA concentrations in wastewater were measured using the EPISENS-S (Efficient and Practical virus Identification System with ENhanced Sensitivity for Solids) method (Ando et al., 2022), with a theoretical limit of detection (LOD) of 93 copies/L. Briefly, solids in each wastewater sample were recovered via low-speed centrifugation, followed by direct RNA extraction from the pellet, multiplex one-step RT-preamplification, and quantitative PCR. Details of the EPISENS-S method were described in a previous study (Ando et al., 2022). To eliminate the effects of dilution by rainwater, the obtained wastewater concentrations were corrected using values adjusted for flow rate to the wastewater treatment plant. This same correction was used in a previous study (Kagami et al., 2025).

### 2.3. Monitoring confirmed COVID-19 cases and hospital testing rates

Data on individuals infected with COVID-19—including age, sex, case category (outpatient, hospital staff member, those infected outside the hospital but developed symptoms after admission, or hospital-acquired infection), and date of case ascertainment— and the number of tests performed each week were extracted from reports by the Department of Infection Control and Prevention at Hokkaido University Hospital. The categories of outpatient, hospital staff member, and those infected outside the hospital but developed symptoms after admission were consolidated into a single category, community-acquired infections. Testing data were then classified into one of two groups, community-or hospital-acquired infection. Testing for community-acquired infections among hospital staff members was partially conducted prior to COVID-19’s reclassification, but not after its reclassification. Therefore, as a sensitivity analysis, we also analyzed community-acquired infections excluding staff members.

### 2.4. Statistical analysis

For SARS-CoV-2 RNA concentrations in wastewater and the cases of community-acquired infections, total infections, and community-acquired infections excluding staff members, we examined how non-detected data were replaced and whether these data were close to normal or log-normal distribution. Among the 2,213 data points of SARS-CoV-2 concentration in wastewater, 249 were below the LOD (93 copies/L). Among the 204-weekly data of community-acquired infection, 28 were below the minimum reported case (MRC; 1 person/week). Similarly, 27 were below the MRC (1 person/week) among the 204 data of total infection cases. The community-acquired infections excluding staff members showed < 1 person/week in 51 out of 204 data. Data replacement for values below the LOD or MRC was performed in accordance with the protocols of a previous study (Murakami et al., 2024) using the value corresponding to half of the non-detected rate based the distribution estimation for left-censored data. This was conducted for both normal and log-normal distributions. For SARS-CoV-2 RNA concentrations in wastewater, community-acquired infection cases, total infection cases, and community-acquired infection cases excluding staff members, we assumed a normal distribution resulted in negative values, and that a log-normal distribution would result in values slightly higher than the LOD or MRC, except for community-acquired infection cases excluding staff members (Table S1). Therefore, as Q-Q plots are effective for evaluating normality when a sample size is large (Mishra et al., 2019), these were created using the following replacements: 0 for a normal distribution and the LOD or MRC value for a log-normal distribution, except community-acquired infection cases excluding staff members.

Regarding community-acquired infection cases excluding staff members, the value corresponding to half of the non-detected rate was used for the for a log-normal distribution. All four variables—SARS-CoV-2 RNA concentration in wastewater, community-acquired infection cases, total infection cases, and community-acquired infection cases excluding staff members—were closer to a log-normal distribution than to a normal distribution (Figure S2). Therefore, in the following analysis, except for community-acquired infection cases excluding staff members, non-detected data were replaced with the LOD or MRC, and log_10_ values were used for SARS-CoV-2 RNA concentrations in wastewater, community-acquired infection cases, and total infection cases.

Regarding community-acquired infection cases excluding staff members, non-detected data were replaced with the value corresponding to half of the non-detected rate, and log_10_ values were used.

Next, linear regression analyses were performed with the log_10_ values of community-acquired infection and total infection cases as the dependent variables and the log_10_ values of SARS-CoV-2 RNA concentrations in wastewater as the independent variable. The representative value of SARS-CoV-2 RNA concentrations in wastewater was calculated as the geometric mean of the measured values obtained during one week, following the protocols used in a previous study (Murakami et al., 2024). A positive correlation was observed between COVID-19 cases and SARS-CoV-2 RNA concentrations in wastewater taken from the same target site between February 2021 and February 2023, with the highest correlation observed when the lag time was set to 0 weeks (Kagami et al., 2023). Therefore, we used the values of wastewater concentration in the same week as the community-acquired infection and total infection cases. To evaluate the impact of changes of in testing rates on the discrepancy between wastewater SARS-CoV-2 RNA concentrations and confirmed cases after the disease’s reclassification to Category LJ, we divided the community-acquired infection and total infection cases by the ratio of the number of clinical tests performed in the corresponding week to determine the average number of clinical tests per week before the reclassification, and then used its log_10_ value as the dependent variables. The testing rate for community-acquired infections was used for those cases, while the total number of tests administered was used for total infection numbers. As described above, the testing rate for community-acquired infections among hospital staff members was not fully ascertained. In this analysis, we assumed that the trend of testing within the hospital represented that of the region as a whole. To complement the assumption, as a sensitive analysis, linear regression analyses were performed with the log_10_ values of community-acquired infections, excluding staff members, as the dependent variables and the log_10_ value of SARS-CoV-2 RNA concentrations in wastewater as the independent variable. For the correction, the clinical testing rate for community-acquired infections excluding staff members was used instead.

Furthermore, we analyzed whether the new cases among hospital-acquired infections could be explained using SARS-CoV-2 RNA wastewater concentration data collected the same week. We defined weeks without incidence among hospital-acquired infections as “0,” weeks with new infections (i.e., the week when a new hospital-acquired infection was acquired) as “1,” and excluded weeks with occurrences among hospital-acquired infections in both the previous and corresponding weeks. A total of 137 weeks were included, of which 22 weeks (16%) had a new case of hospital-acquired infection. We performed a modified Poisson regression analysis (i.e., Poisson regression analysis with a robust error variance) with the new incidence among hospital-acquired infections as the dependent variable and the log_10_ value of the SARS-CoV-2 RNA concentration in wastewater as the independent variable. The modified Poisson regression analysis was chosen because, even when the prevalence was 10% or higher, the incidence rate ratio it calculated was close to the relative risk (McNutt et al., 2003; Zou, 2004). Based on the partial regression coefficients obtained, we calculated the log_10_ value of SARS-CoV-2 RNA concentration in wastewater (95% confidence interval [CI]) corresponding to a 25% probability of a new incidence. The value of 25% was chosen because it is slightly higher than the 16% prevalence of the new incidence rate observed throughout the survey period in this study, making it an appropriate threshold for alert.

Finally, we analyzed the association of the log_10_ value of the SARS-CoV-2 RNA concentration in wastewater at the start of the new incidence with the number of continuous weeks from the start through the end of new incidence among hospital-acquired infections (hereinafter referred to as an “event”) and the total number of hospital-acquired infections during the event. After using Q-Q plots to confirm that both the number of continuous weeks and the total number of hospital-acquired infections per event were closer to a log-normal distribution than to a normal distribution (Figure S3), we performed linear regression analyses with their log_10_ values as the dependent variables and the log_10_ values of SARS-CoV-2 RNA concentration in wastewater during the week when the new incidence among hospital-acquired infections appeared as the independent variable.

The analysis was performed using R (R Development Core Team, 2025); R packages “NADA” (Lee, 2022), “sandwich” (The Comprehensive R Archive Network, 2025b), and “lmtest” (The Comprehensive R Archive Network, 2025a); and IBM SPSS version 28.

## 3. Results

Figure 1(a–c) and Table S2 show the temporal changes in SARS-CoV-2 RNA concentrations in wastewater in the City of Sapporo, the confirmed COVID-19 cases, and the number tests conducted at Hokkaido University Hospital. The number of confirmed COVID-19 cases corrected by the number of tests performed are also shown in Figure 1(d). Table 1 presents the characteristics of community-acquired infections, hospital-acquired infections, total infections, and the ages of those infected before and after COVID-19’s reclassification to Category LJ. In general, SARS-CoV-2 RNA concentrations in wastewater remained high even after the reclassification, but the numbers of community-acquired infections and total infection cases peaked before the reclassification. The number of tests conducted decreased considerably after the reclassification. Significant positive associations were observed between SARS-CoV-2 RNA concentrations in wastewater and the numbers of community-acquired and total infections (Figures 2a–d). Without correcting based on the testing rate, the *R*² values were 0.54 (*P* < 0.001) for community-acquired infections and 0.57 (*P* < 0.001) for all infections. Infection cases after the reclassification were lower than those estimated using the regression equations. In contrast, after correcting for the testing rate, the *R*² values improved to 0.75 (*P* < 0.001) for community-acquired infections and 0.78 (*P* < 0.001) for all cases (Figure 2c, d). The slopes with correction using the testing rate were 0.85 (95% CI: 0.78–0.91) for community-acquired infections and 0.66 (95% CI: 0.62–0.71) for all cases, both of which were significantly lower than 1. A similar result was obtained for community-acquired infections excluding staff members (Figure S4). The *R*² value was 0.44 (*P* < 0.001) for non-corrected data based on the testing rate and 0.62 (*P* < 0.001) for corrections accounting for the number of tests conducted. The slope with correction was 0.67 (95% CI: 0.60–0.74).

**Figure 1.**
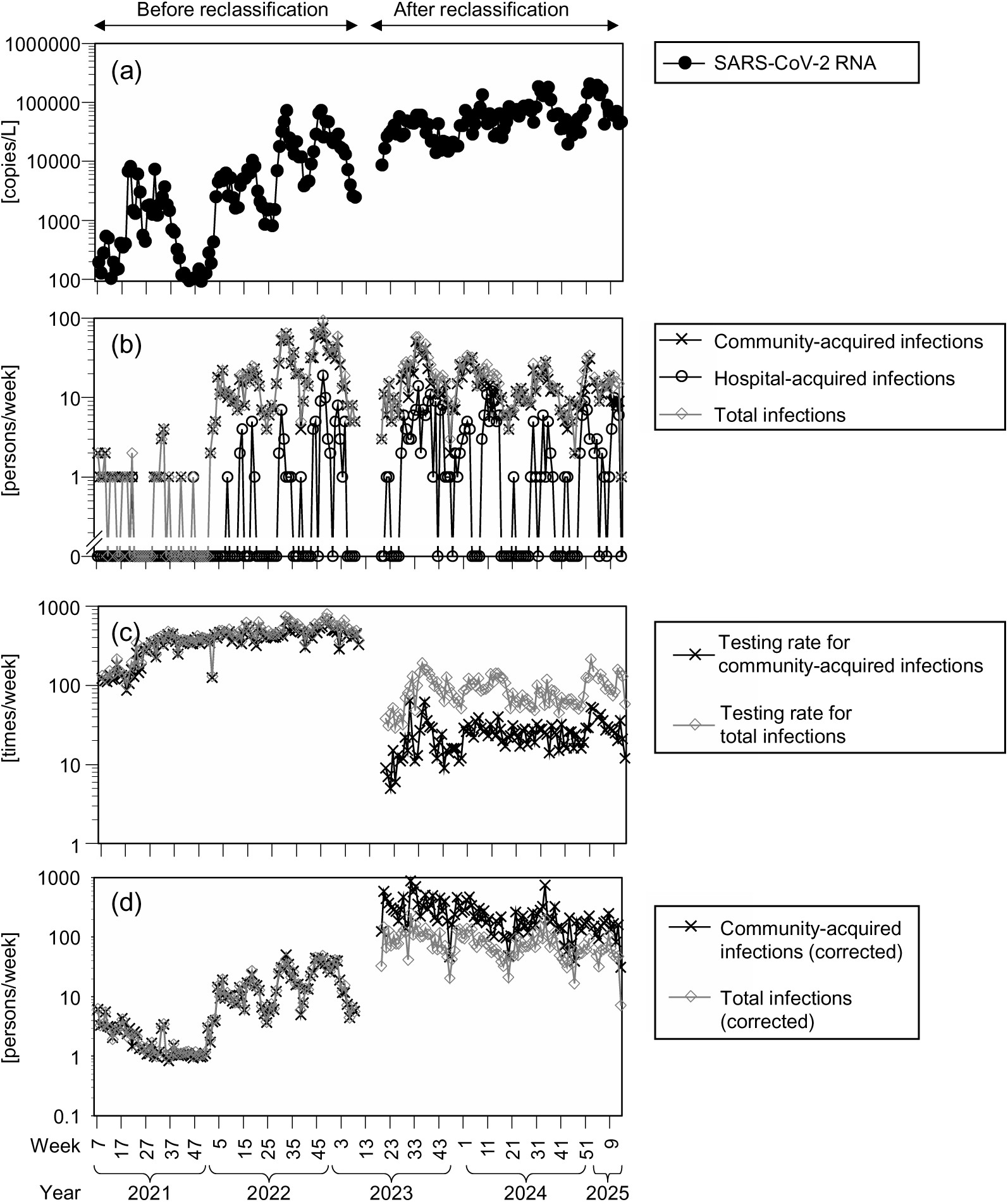
Temporal changes in SARS-CoV-2 RNA concentrations in wastewater, confirmed COVID-19 cases, and the testing rate. (a) Wastewater concentrations in the City of Sapporo, (b) Confirmed COVID-19 cases at Hokkaido University Hospital (without correction for testing rate), (c) Hospital testing rate, (d) Confirmed COVID-19 cases at the hospital (with correction for testing rate).

**Figure 2.**
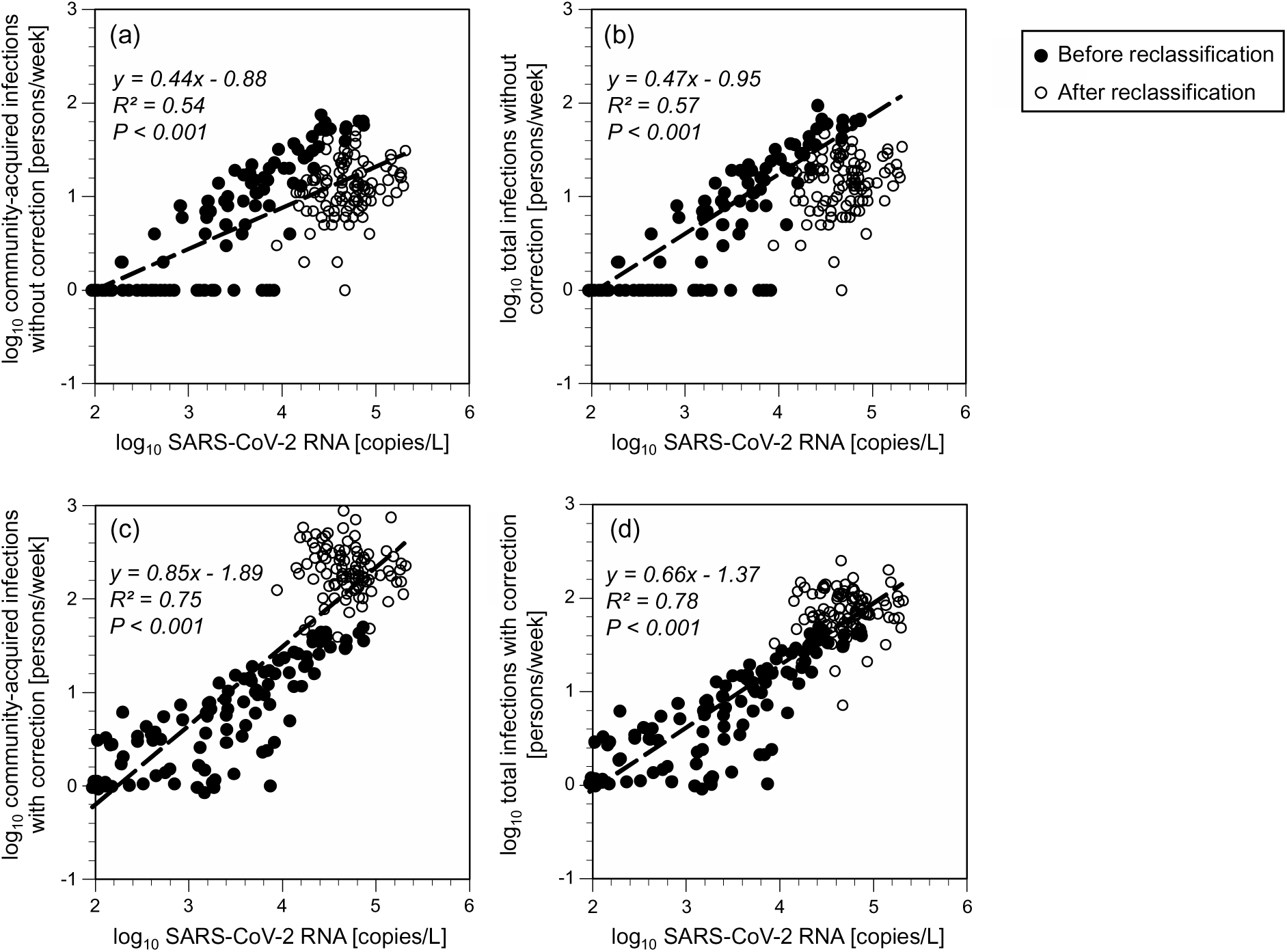
Scatterplots of SARS-CoV-2 RNA concentration vs. confirmed COVID-19 cases. (a) Community-acquired infections without correction by the number of testing, (b) Total infections without correction for testing rate, (c) Community-acquired infections with correction for testing rate, (d) Total infections with correction for testing rate.

**Table 1.** Characteristics of individuals infected with COVID-19.

|  |  | Male |  |  | Female |  |  | Total |  |  |
| --- | --- | --- | --- | --- | --- | --- | --- | --- | --- | --- |
|  |  | *N* | Age (arithmetic mean) | Age (median) | *N* | Age (arithmetic mean) | Age (median) | *N* | Age (arithmetic mean) | Age (median) |
| Before classification | Community-acquired infections | 567 | 42 | 38 | 836 | 40 | 37 | 1403 | 41 | 37 |
|  | Hospital-acquired infections | 63 | 58 | 66 | 42 | 56 | 62 | 105 | 57 | 62 |
|  | Total infections | 630 | 43 | 39 | 878 | 40 | 37 | 1508 | 42 | 38 |
| After classification | Community-acquired infections | 538 | 46 | 41 | 877 | 41 | 38 | 1415 | 43 | 39 |
|  | Hospital-acquired infections | 151 | 61 | 66 | 127 | 61 | 66 | 278 | 61 | 66 |
|  | Total infections | 689 | 49 | 47 | 1004 | 44 | 40 | 1693 | 46 | 43 |

A significant positive association was observed between the SARS-CoV-2 RNA concentration in wastewater and the new incidences of hospital-acquired infections (Figure 3). The incidence risk rate of log_10_ SARS-CoV-2 RNA concentration based on new incidences among hospital-acquired infections was 2.24 (95% CI: 1.36–3.71) (*P* = 0.002), and the log_10_ for SARS-CoV-2 RNA concentration [copies/L] at which there was a 25% probability of a new incidence was 4.57 (95% CI: 4.10–5.03). The SARS-CoV-2 RNA concentration in wastewater at the onset of a new case showed a weak, but significantly positive, association with the number of continuous weeks and the total number of hospital-acquired infections per event (*R*² = 0.26, *P* = 0.016; *R*² = 0.23, *P* = 0.024, respectively) (Figure 4).

**Figure 3.**
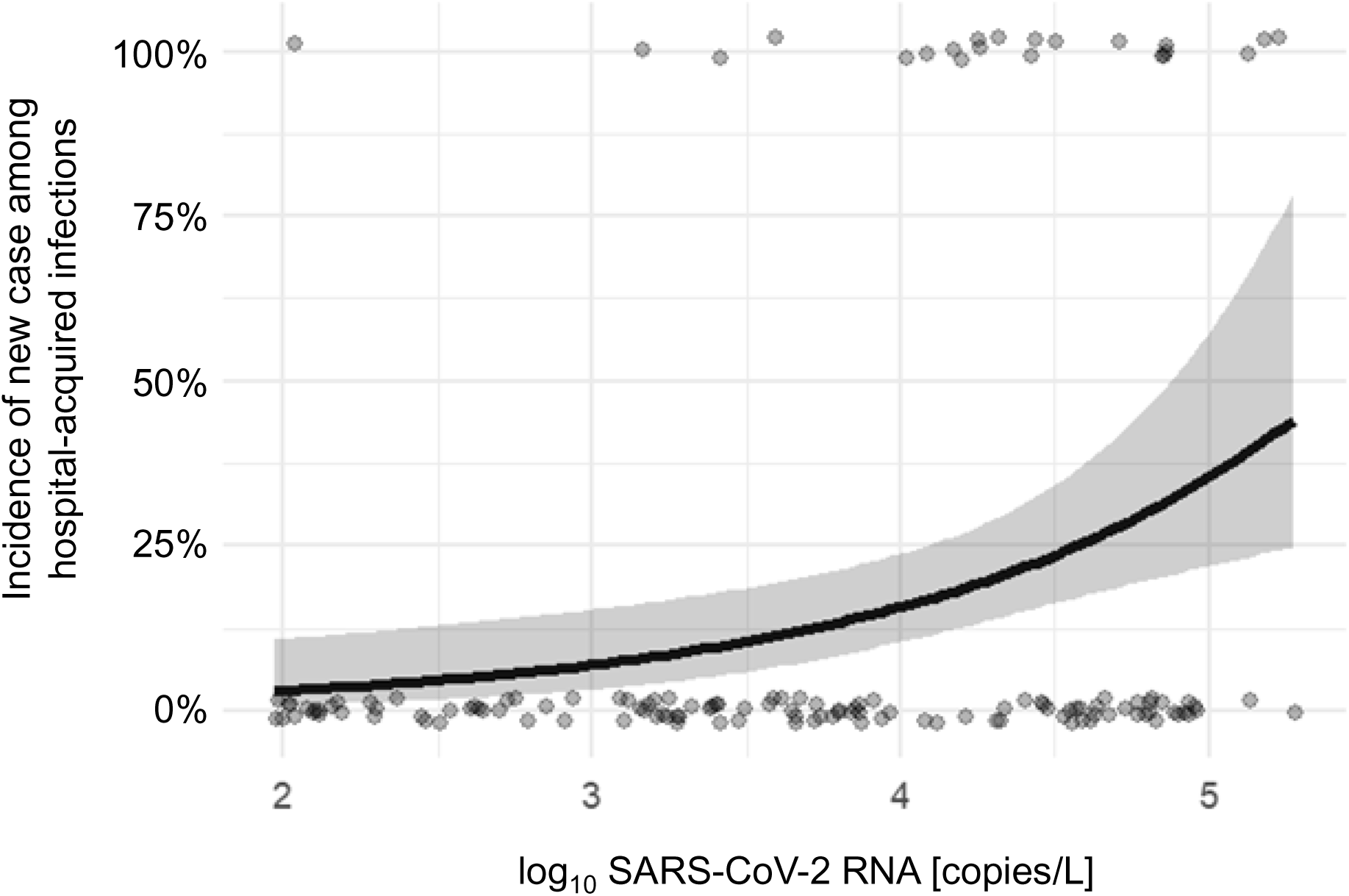
Associations between SARS-CoV-2 RNA concentrations in wastewater and the incidence of new cases among hospital-acquired infections. Solid line: modified Poisson regression analysis was applied in the prediction. Gray areas represent a 95% confidence interval (CI) of the prediction. Partial regression coefficient (95% CI): intercept, −5.08 (−7.30–−2.85) (*P* < 0.001); log_10_ SARS-CoV-2 RNA concentrations, 0.81 (0.31–1.31) (*P* = 0.002).

**Figure 4.**
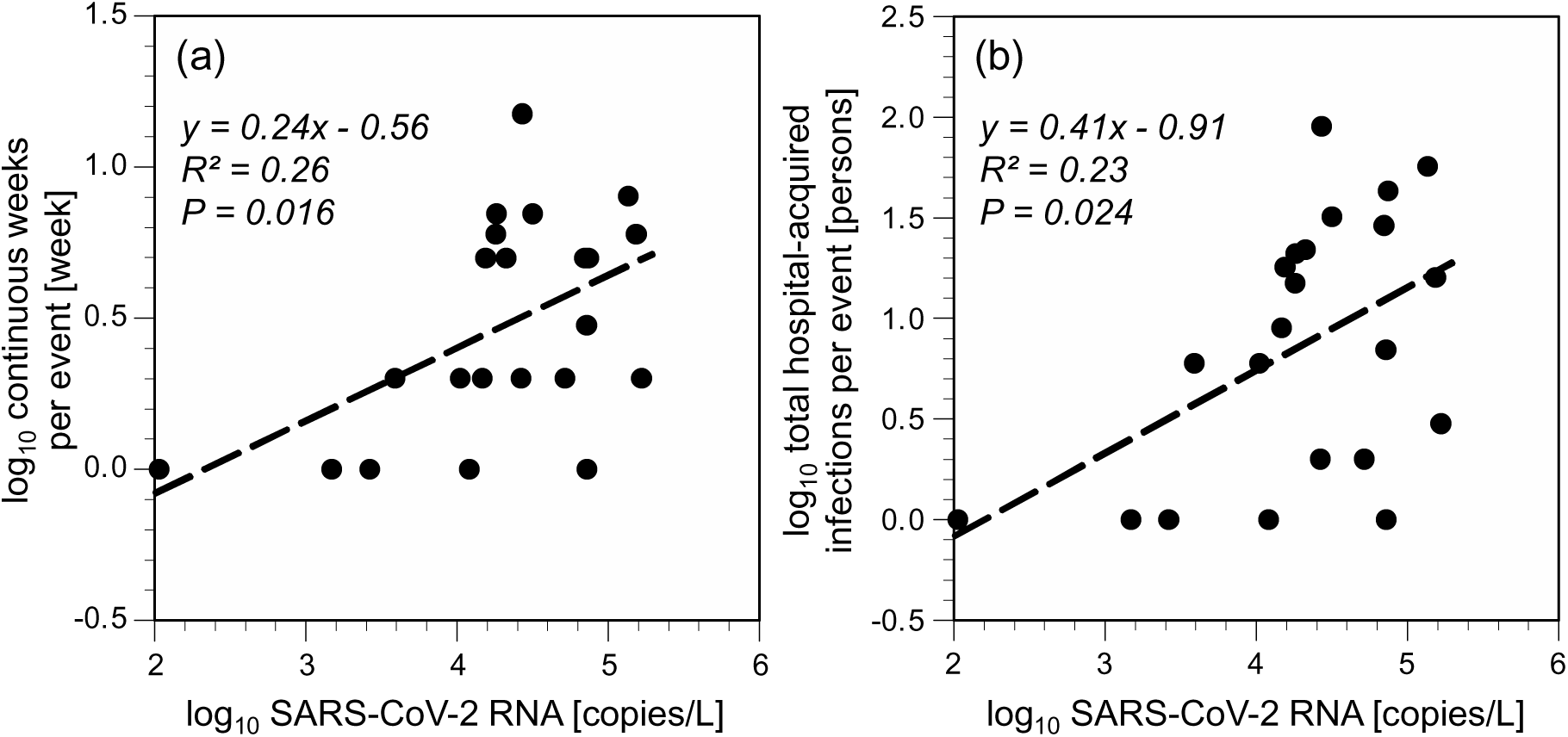
Associations of SARS-CoV-2 RNA concentration at the start of the new incidence with continuous weeks or total hospital-acquired infections per event. (a) Continuous weeks, (b) Total hospital-acquired infections.

## 4. Discussion

This study investigated the association between SARS-CoV-2 RNA concentrations in wastewater samples collected in Sapporo, Japan with confirmed cases of COVID-19 at Hokkaido University Hospital over a period of approximately four years. With the disease’s reclassification to Category LJ, the testing rate decreased considerably. After correcting for this, strong associations were demonstrated between SARS-CoV-2 RNA concentrations in wastewater and cases of community-acquired infection or total infections at the hospital. This was also confirmed by a sensitivity analysis that excluded staff members. In the same study site, we found that a strong correlation between SARS-CoV-2 RNA concentrations in wastewater and the confirmed number of COVID-19 cases was reported before the reclassification (Kagami et al., 2023), while the infection cases per SARS-CoV-2 RNA concentration in wastewater was reduced after the reclassification (Kagami et al., 2025). These results are consistent with the findings of previous studies (Kagami et al., 2023; Kagami et al., 2025) and provided new insights suggesting that COVID-19 cases at the hospital could be sufficiently explained by SARS-CoV-2 RNA concentrations in wastewater, after correcting for the testing rate. Therefore, the divergence between SARS-CoV-2 RNA concentrations in wastewater and COVID-19 cases after the reclassification was largely attributable to a decrease in the number tests conducted (Boehm et al., 2023). This highlights that SARS-CoV-2 wastewater surveillance is an objective indicator that reflects infection incidence, including asymptomatic infections, without being influenced by factors such as changes in hospital visiting behaviors and the number of tests conducted.

This study has also demonstrated that new cases among hospital-acquired infections were associated with SARS-CoV-2 RNA concentrations in wastewater. Considering that SARS-CoV-2 RNA concentrations in wastewater reflect community infection incidence, this suggests that hospital-acquired infections might occur through visits to hospitals by infected patients, hospital staff members, or members of the community (Hatfield et al., 2023; Kagami et al., 2025). Moreover, the positive associations between SARS-CoV-2 RNA concentrations in wastewater with the number of continuous weeks and the total number of hospital-acquired infections per event suggest that higher concentrations were associated with longer-lasting and large numbers of incidences among hospital-acquired infections. Therefore, wastewater surveillance is expected to serve as a valid indicator regarding new incidences among hospital-acquired infections. This study found that a 25% probability of a new incidence corresponded to a log_10_ SARS-CoV-2 RNA concentration [copies/L] of 4.57 (95% CI: 4.10–5.03).

This study has several limitations. First, it was focused only on the association between SARS-CoV-2 RNA concentrations in wastewater in Sapporo and the confirmed number COVID-19 cases at Hokkaido University Hospital, so caution is needed when extending these findings to other regions.

Second, while the study demonstrated that COVID-19 cases at the hospital could be explained via wastewater SARS-CoV-2 RNA concentrations after correcting for the testing rate, the reasons for the decline in the number of tests conducted after the disease’s reclassification remain unclear. One possible interpretation is that individuals avoided hospital visits, due to the introduction of testing or therapy fees after the reclassification to Category LJ (Maree et al., 2025). However, other factors, including an increase in asymptomatic or mild cases due to viral attenuation (Nakakubo et al., 2023), cannot be ruled out.

Third, we could not completely ascertain the testing rate for community-acquired infections among staff members within the hospital. Therefore, in this study, a sensitivity analysis was performed by excluding staff members infected with COVID-19. When we did this, we could confirm that consistent results were obtained.

Fourth, this study did not adequately consider factors regarding the hospital’s capacity for individuals infected with COVID-19 or visits to other hospitals. In particular, regressions between the log_10_ values of SARS-CoV-2 RNA concentrations in wastewater and the log_10_ values of community-acquired or total infection cases showed that their slopes were less than 1. In a log–log regression equation, a slope of 1 indicates that the two variables are proportional (Y = a × X^b^ _¢:_:> log_10_ Y = b × log_10_ X + log_10_ a). Therefore, wastewater virus concentrations were not proportional to the infected individual cases at the hospital. This result contrasts with a previous study that showed the slope between the log_10_ values of SARS-CoV-2 RNA concentrations in wastewater and the log_10_ values of COVID-19 cases in Sapporo was nearly 1 (Murakami et al., 2024). One possible interpretation is that visits to other hospitals might be a factor; therefore, a comprehensive survey targeting individuals infected with COVID-19 across multiple hospital facilities is a promising area for future research.

## 5. Conclusions

This study analyzed the associations between SARS-CoV-2 RNA concentrations in wastewater and confirmed COVID-19 cases at a university hospital, with and without correction accounting for testing conducted within the hospital, to clarify the cause of the discrepancy between wastewater viral concentrations and confirmed cases after COVID’s reclassification to Category LJ. By correcting for the number of tests conducted at the hospital, the community-acquired infections and total number of infection cases at the hospital could be explained by the SARS-CoV-2 RNA concentration in wastewater, and the discrepancy was interpreted as being mainly due to a decrease in the testing rate.

This study emphasized that SARS-CoV-2 wastewater surveillance is an objective indicator reflecting infection incidence that is independent of individuals’ hospital visiting behaviors and testing rates.

Furthermore, SARS-CoV-2 RNA concentrations in wastewater were positively associated with the probability and total number of new cases among hospital-acquired infections and their continuous time length. As an indicator for alerting to incidence among hospital-acquired infections, a log_10_ SARS-CoV-2 RNA concentration [copies/L] of 4.57 (95% CI: 4.10–5.03) in wastewater is suggested as a 25% probability of new incidence.

## Data Availability

All data produced in the present study are available upon reasonable request to the authors.

## Acknowledgement

We acknowledge the Business Quest Co., Ltd. (www.speedtensaku.com/) for English language editing.

## Conflicts of Interest

Michio Murakami reports a relationship with NJS Co., Ltd. that includes: consulting or advisory. Masaaki Kitajima received funding grants and speaking fees from Shionogi & Co., Ltd. and AdvanSentinel, Inc. and funding grants from Shimadzu Corporation, and he has a patent with royalties paid from Shionogi and Co. Ltd. Other authors declare no competing financial interest.

## Funding

This work was supported by “The Nippon Foundation-Osaka University Project for Infectious Disease Prevention,” the MHLW Research on Health Security Control Program, under grant number JPMH2024LA1006, the Japan Science and Technology Agency (JST) through the JST-Mirai Program, under grant JPMJMI22D1 and the Japan Agency for Medical Research and Development (AMED), under grant 24fk0108713h0001.

## Notes

### Funding Statement

This study was funded by “The Nippon Foundation-Osaka University Project for Infectious Disease Prevention,” the MHLW Research on Health Security Control Program, under grant number JPMH2024LA1006, the Japan Science and Technology Agency (JST) through the JST-Mirai Program, under grant JPMJMI22D1 and the Japan Agency for Medical Research and Development (AMED), under grant 24fk0108713h0001.

### Author Declarations

The Institutional Review Board of Hokkaido University Hospital for Clinical Research gave ethical approval for this work (approval number: 025-0012).

