## Supplementary material for "Wastewater Surveillance Reveals Testing-Related Underreporting and Hospital-Acquired SARS-CoV-2 Infections": Tables S1 and S2; Figure S1-4.

Table S1. Value corresponding to half of the ND (non-detected) rate.

| Items | Distribution | Value corresponding to half of the ND rate |
|---|---|---|
| SARS-CoV-2 RNA concentrations in wastewater [copies/L] | Normal distribution | −83657 |
| SARS-CoV-2 RNA concentrations in wastewater [copies/L] | Log-normal distribution | 99.7 |
| Community-acquired infections [persons/week] | Normal distribution | −7.53 |
| Community-acquired infections [persons/week] | Log-normal distribution | 1.18 |
| Total infections [persons/week] | Normal distribution | −8.65 |
| Total infections [persons/week] | Log-normal distribution | 1.18 |
| Community-acquired infections excluding staff members [persons/week | Normal distribution | −0.34 |
| Community-acquired infections excluding staff members [persons/week | Log-normal distribution | 0.95 |

Table S2. SARS-CoV-2 RNA concentrations in wastewater, confirmed COVID-19 cases, and the testing rate for each week.

| Year | Week | The number of wastewater samples [samples/ week] | SARS-CoV-2 RNA [copies/L] | Community -acquired infections [persons/ week] | Hospital-acquired infections [persons/ week] | Total infections [persons/ week] | Community -acquired infections excluding staff members [persons/ week] | Testing rate for community-acquired infections [times/ week] | Testing rate for total infections [times/ week] | Testing rate for community-acquired infections excluding staff members [times/ week] |
|---|---|---|---|---|---|---|---|---|---|---|
| 2021 | 07 | 9 | 196 | 2 | 0 | 2 | 1 | 122 | 133 | 116 |
| 2021 | 08 | 9 | 129 | 1 | 0 | 1 | 0 | 114 | 126 | 111 |
| 2021 | 09 | 9 | 282 | 1 | 0 | 1 | 0 | 110 | 120 | 106 |
| 2021 | 10 | 9 | 537 | 2 | 0 | 2 | 1 | 135 | 152 | 131 |
| 2021 | 11 | 9 | 500 | 0 | 0 | 0 | 0 | 118 | 137 | 110 |
| 2021 | 12 | 9 | 105 | 1 | 0 | 1 | 1 | 121 | 143 | 115 |
| 2021 | 13 | 9 | 198 | 1 | 0 | 1 | 0 | 182 | 215 | 169 |
| 2021 | 14 | 9 | 144 | 1 | 0 | 1 | 0 | 135 | 154 | 129 |
| 2021 | 15 | 15 | 151 | 0 | 0 | 0 | 0 | 134 | 143 | 124 |
| 2021 | 16 | 15 | 412 | 0 | 0 | 0 | 0 | 121 | 134 | 112 |
| 2021 | 17 | 15 | 351 | 1 | 0 | 1 | 0 | 86 | 100 | 73 |
| 2021 | 18 | 9 | 399 | 1 | 0 | 1 | 0 | 105 | 131 | 101 |
| 2021 | 19 | 15 | 6828 | 1 | 0 | 1 | 0 | 157 | 195 | 141 |
| 2021 | 20 | 15 | 8175 | 0 | 0 | 0 | 0 | 128 | 171 | 118 |
| 2021 | 21 | 15 | 1481 | 1 | 1 | 2 | 1 | 254 | 344 | 246 |
| 2021 | 22 | 15 | 1318 | 0 | 0 | 0 | 0 | 145 | 184 | 134 |
| 2021 | 23 | 15 | 6110 | 0 | 0 | 0 | 0 | 163 | 194 | 158 |
| 2021 | 24 | 15 | 3039 | 0 | 0 | 0 | 0 | 278 | 298 | 268 |
| 2021 | 25 | 15 | 560 | 0 | 0 | 0 | 0 | 269 | 281 | 263 |
| 2021 | 26 | 15 | 444 | 0 | 0 | 0 | 0 | 290 | 304 | 279 |
| 2021 | 27 | 15 | 1775 | 0 | 0 | 0 | 0 | 344 | 357 | 334 |
| 2021 | 28 | 15 | 1859 | 0 | 0 | 0 | 0 | 338 | 356 | 330 |
| 2021 | 29 | 15 | 1274 | 0 | 0 | 0 | 0 | 226 | 245 | 224 |
| 2021 | 30 | 15 | 7401 | 1 | 0 | 1 | 0 | 372 | 398 | 369 |
| 2021 | 31 | 15 | 1228 | 1 | 0 | 1 | 0 | 387 | 419 | 380 |
| 2021 | 32 | 15 | 1900 | 1 | 0 | 1 | 1 | 320 | 337 | 309 |
| 2021 | 33 | 15 | 2519 | 3 | 0 | 3 | 0 | 386 | 401 | 363 |
| 2021 | 34 | 15 | 3719 | 4 | 0 | 4 | 0 | 437 | 478 | 417 |
| 2021 | 35 | 15 | 1858 | 0 | 0 | 0 | 0 | 390 | 404 | 379 |
| 2021 | 36 | 15 | 1466 | 1 | 0 | 1 | 0 | 443 | 455 | 436 |
| 2021 | 37 | 15 | 701 | 0 | 0 | 0 | 0 | 357 | 378 | 350 |

| | | | | | | | | | | |
|---|---|---|---|---|---|---|---|---|---|---|
| 2021 | 38 | 15 | 628 | 0 | 0 | 0 | 0 | 247 | 260 | 237 |
| 2021 | 39 | 15 | 323 | 0 | 0 | 0 | 0 | 354 | 369 | 350 |
| 2021 | 40 | 15 | 231 | 0 | 0 | 0 | 0 | 367 | 379 | 354 |
| 2021 | 41 | 15 | 120 | 1 | 0 | 1 | 1 | 362 | 374 | 349 |
| 2021 | 42 | 15 | 128 | 0 | 0 | 0 | 0 | 351 | 363 | 344 |
| 2021 | 43 | 15 | 107 | 0 | 0 | 0 | 0 | 340 | 351 | 336 |
| 2021 | 44 | 15 | 96 | 0 | 0 | 0 | 0 | 335 | 345 | 326 |
| 2021 | 45 | 15 | 101 | 0 | 0 | 0 | 0 | 374 | 380 | 367 |
| 2021 | 46 | 15 | 106 | 0 | 1 | 1 | 0 | 401 | 420 | 391 |
| 2021 | 47 | 15 | 104 | 0 | 0 | 0 | 0 | 333 | 352 | 331 |
| 2021 | 48 | 15 | 150 | 0 | 0 | 0 | 0 | 380 | 399 | 373 |
| 2021 | 49 | 15 | 93 | 0 | 0 | 0 | 0 | 386 | 395 | 379 |
| 2021 | 50 | 15 | 126 | 0 | 0 | 0 | 0 | 373 | 383 | 368 |
| 2021 | 51 | 15 | 128 | 0 | 0 | 0 | 0 | 343 | 358 | 333 |
| 2021 | 52 | 10 | 282 | 0 | 0 | 0 | 0 | 125 | 130 | 118 |
| 2022 | 01 | 15 | 190 | 2 | 0 | 2 | 1 | 436 | 445 | 430 |
| 2022 | 02 | 15 | 433 | 4 | 0 | 4 | 2 | 395 | 410 | 382 |
| 2022 | 03 | 15 | 2531 | 5 | 0 | 5 | 0 | 467 | 489 | 445 |
| 2022 | 04 | 15 | 4505 | 18 | 0 | 18 | 0 | 465 | 493 | 428 |
| 2022 | 05 | 15 | 5445 | 11 | 0 | 11 | 3 | 436 | 455 | 400 |
| 2022 | 06 | 15 | 4756 | 22 | 0 | 22 | 8 | 431 | 469 | 405 |
| 2022 | 07 | 15 | 6369 | 12 | 0 | 12 | 4 | 474 | 509 | 440 |
| 2022 | 08 | 15 | 2635 | 10 | 1 | 11 | 3 | 360 | 394 | 340 |
| 2022 | 09 | 15 | 5265 | 11 | 0 | 11 | 6 | 389 | 425 | 368 |
| 2022 | 10 | 15 | 2446 | 9 | 0 | 9 | 1 | 398 | 419 | 379 |
| 2022 | 11 | 15 | 1616 | 9 | 0 | 9 | 1 | 438 | 474 | 424 |
| 2022 | 12 | 15 | 1664 | 7 | 0 | 7 | 2 | 335 | 358 | 326 |
| 2022 | 13 | 15 | 3888 | 17 | 2 | 19 | 4 | 451 | 533 | 432 |
| 2022 | 14 | 15 | 5141 | 15 | 4 | 19 | 1 | 558 | 622 | 541 |
| 2022 | 15 | 15 | 5154 | 8 | 0 | 8 | 3 | 500 | 531 | 482 |
| 2022 | 16 | 15 | 7252 | 20 | 0 | 20 | 1 | 438 | 472 | 414 |
| 2022 | 17 | 15 | 6384 | 16 | 0 | 16 | 3 | 361 | 402 | 331 |
| 2022 | 18 | 9 | 10447 | 20 | 5 | 25 | 4 | 316 | 379 | 299 |
| 2022 | 19 | 15 | 8272 | 23 | 1 | 24 | 4 | 541 | 626 | 533 |
| 2022 | 20 | 15 | 3143 | 19 | 0 | 19 | 1 | 466 | 534 | 443 |
| 2022 | 21 | 15 | 2090 | 14 | 0 | 14 | 1 | 416 | 454 | 391 |
| 2022 | 22 | 15 | 1720 | 7 | 0 | 7 | 0 | 393 | 420 | 384 |
| 2022 | 23 | 15 | 856 | 6 | 0 | 6 | 1 | 437 | 485 | 415 |
| 2022 | 24 | 15 | 1504 | 4 | 0 | 4 | 0 | 408 | 444 | 394 |
| 2022 | 25 | 15 | 1567 | 6 | 0 | 6 | 2 | 400 | 439 | 382 |

| | | | | | | | | | | |
|---|---|---|---|---|---|---|---|---|---|---|
| 2022 | 26 | 15 | 817 | 8 | 0 | 8 | 0 | 407 | 443 | 395 |
| 2022 | 27 | 15 | 1534 | 7 | 0 | 7 | 1 | 430 | 465 | 415 |
| 2022 | 28 | 15 | 7034 | 15 | 0 | 15 | 6 | 458 | 495 | 435 |
| 2022 | 29 | 15 | 18043 | 27 | 2 | 29 | 3 | 416 | 453 | 387 |
| 2022 | 30 | 15 | 32454 | 53 | 7 | 60 | 12 | 649 | 747 | 585 |
| 2022 | 31 | 15 | 47756 | 52 | 3 | 55 | 8 | 634 | 713 | 568 |
| 2022 | 32 | 15 | 72793 | 64 | 1 | 65 | 13 | 477 | 630 | 420 |
| 2022 | 33 | 15 | 25289 | 52 | 1 | 53 | 6 | 561 | 625 | 496 |
| 2022 | 34 | 15 | 18479 | 27 | 1 | 28 | 3 | 494 | 551 | 438 |
| 2022 | 35 | 15 | 13309 | 37 | 0 | 37 | 11 | 513 | 596 | 474 |
| 2022 | 36 | 15 | 21820 | 20 | 0 | 20 | 3 | 472 | 512 | 445 |
| 2022 | 37 | 15 | 11865 | 20 | 0 | 20 | 4 | 461 | 513 | 430 |
| 2022 | 38 | 15 | 12024 | 4 | 1 | 5 | 0 | 300 | 349 | 285 |
| 2022 | 39 | 15 | 3851 | 9 | 0 | 9 | 4 | 424 | 472 | 406 |
| 2022 | 40 | 15 | 4392 | 17 | 0 | 17 | 1 | 462 | 513 | 437 |
| 2022 | 41 | 15 | 4672 | 14 | 0 | 14 | 3 | 393 | 442 | 374 |
| 2022 | 42 | 15 | 9124 | 32 | 0 | 32 | 5 | 540 | 591 | 508 |
| 2022 | 43 | 15 | 14692 | 32 | 4 | 36 | 5 | 461 | 510 | 423 |
| 2022 | 44 | 15 | 28856 | 62 | 5 | 67 | 8 | 527 | 608 | 452 |
| 2022 | 45 | 15 | 65733 | 64 | 0 | 64 | 13 | 552 | 635 | 484 |
| 2022 | 46 | 15 | 74009 | 58 | 9 | 67 | 18 | 609 | 704 | 568 |
| 2022 | 47 | 15 | 25935 | 75 | 19 | 94 | 11 | 634 | 793 | 551 |
| 2022 | 48 | 15 | 48200 | 56 | 10 | 66 | 10 | 582 | 685 | 521 |
| 2022 | 49 | 15 | 46859 | 39 | 3 | 42 | 6 | 491 | 575 | 451 |
| 2022 | 50 | 15 | 24466 | 34 | 2 | 36 | 4 | 494 | 572 | 449 |
| 2022 | 51 | 15 | 20971 | 44 | 0 | 44 | 11 | 471 | 566 | 432 |
| 2022 | 52 | 10 | 21136 | 31 | 5 | 36 | 3 | 286 | 360 | 246 |
| 2023 | 01 | 10 | 29246 | 51 | 8 | 59 | 7 | 481 | 613 | 437 |
| 2023 | 02 | 15 | 17222 | 26 | 3 | 29 | 10 | 512 | 662 | 483 |
| 2023 | 03 | 15 | 15912 | 13 | 1 | 14 | 2 | 416 | 476 | 399 |
| 2023 | 04 | 15 | 13207 | 14 | 5 | 19 | 3 | 450 | 511 | 426 |
| 2023 | 05 | 15 | 7296 | 8 | 0 | 8 | 2 | 402 | 461 | 386 |
| 2023 | 06 | 15 | 4046 | 5 | 0 | 5 | 1 | 419 | 469 | 406 |
| 2023 | 07 | 15 | 2611 | 8 | 0 | 8 | 2 | 452 | 492 | 436 |
| 2023 | 08 | 15 | 2493 | 5 | 0 | 5 | 2 | 327 | 370 | 316 |
| 2023 | 19 | 15 | 8729 | 3 | 0 | 3 | 0 | 9 | 38 | 9 |
| 2023 | 20 | 15 | 16608 | 11 | 0 | 11 | 3 | 7 | 31 | 7 |
| 2023 | 21 | 15 | 26460 | 6 | 1 | 7 | 2 | 5 | 44 | 5 |
| 2023 | 22 | 15 | 30176 | 15 | 1 | 16 | 2 | 15 | 52 | 15 |
| 2023 | 23 | 15 | 33960 | 5 | 0 | 5 | 1 | 6 | 29 | 6 |

| | | | | | | | | | | |
|---|---|---|---|---|---|---|---|---|---|---|
| 2023 | 24 | 15 | 41536 | 10 | 0 | 10 | 1 | 13 | 40 | 13 |
| 2023 | 25 | 15 | 29261 | 8 | 0 | 8 | 2 | 11 | 43 | 11 |
| 2023 | 26 | 15 | 57319 | 6 | 0 | 6 | 2 | 12 | 32 | 12 |
| 2023 | 27 | 15 | 26957 | 17 | 2 | 19 | 4 | 22 | 70 | 22 |
| 2023 | 28 | 15 | 28843 | 19 | 6 | 25 | 6 | 15 | 80 | 15 |
| 2023 | 29 | 15 | 48578 | 24 | 4 | 28 | 5 | 64 | 97 | 64 |
| 2023 | 30 | 15 | 48094 | 10 | 3 | 13 | 5 | 22 | 130 | 22 |
| 2023 | 31 | 15 | 44926 | 26 | 3 | 29 | 4 | 11 | 48 | 11 |
| 2023 | 32 | 15 | 44526 | 20 | 6 | 26 | 5 | 13 | 99 | 13 |
| 2023 | 33 | 15 | 60483 | 51 | 7 | 58 | 16 | 27 | 146 | 27 |
| 2023 | 34 | 15 | 60668 | 44 | 14 | 58 | 8 | 46 | 191 | 46 |
| 2023 | 35 | 15 | 60581 | 37 | 2 | 39 | 9 | 62 | 159 | 62 |
| 2023 | 36 | 15 | 46273 | 32 | 7 | 39 | 7 | 36 | 156 | 36 |
| 2023 | 37 | 15 | 30610 | 41 | 6 | 47 | 7 | 30 | 138 | 30 |
| 2023 | 38 | 15 | 42114 | 23 | 9 | 32 | 2 | 29 | 119 | 29 |
| 2023 | 39 | 15 | 22162 | 15 | 11 | 26 | 3 | 16 | 97 | 16 |
| 2023 | 40 | 5 | 22634 | 16 | 1 | 17 | 4 | 12 | 110 | 12 |
| 2023 | 41 | 5 | 14087 | 11 | 9 | 20 | 1 | 19 | 89 | 19 |
| 2023 | 42 | 5 | 43916 | 12 | 0 | 12 | 4 | 24 | 81 | 24 |
| 2023 | 43 | 5 | 15447 | 11 | 7 | 18 | 5 | 9 | 63 | 9 |
| 2023 | 44 | 5 | 22200 | 11 | 8 | 19 | 5 | 15 | 126 | 15 |
| 2023 | 45 | 5 | 21129 | 15 | 1 | 16 | 7 | 14 | 93 | 14 |
| 2023 | 46 | 5 | 15055 | 8 | 1 | 9 | 2 | 16 | 80 | 16 |
| 2023 | 47 | 5 | 17146 | 2 | 1 | 3 | 0 | 16 | 61 | 16 |
| 2023 | 48 | 5 | 21376 | 7 | 0 | 7 | 4 | 16 | 69 | 16 |
| 2023 | 49 | 5 | 18223 | 7 | 2 | 9 | 0 | 11 | 63 | 11 |
| 2023 | 50 | 5 | 18310 | 15 | 1 | 16 | 4 | 12 | 51 | 12 |
| 2023 | 51 | 5 | 40488 | 26 | 2 | 28 | 1 | 29 | 86 | 29 |
| 2023 | 52 | 5 | 40676 | 18 | 3 | 21 | 6 | 26 | 89 | 26 |
| 2024 | 01 | 5 | 73651 | 24 | 4 | 28 | 6 | 32 | 122 | 32 |
| 2024 | 02 | 5 | 49820 | 29 | 5 | 34 | 9 | 26 | 101 | 26 |
| 2024 | 03 | 5 | 59547 | 28 | 4 | 32 | 8 | 22 | 107 | 22 |
| 2024 | 04 | 5 | 29308 | 32 | 0 | 32 | 10 | 35 | 98 | 35 |
| 2024 | 05 | 5 | 43195 | 23 | 0 | 23 | 8 | 39 | 123 | 39 |
| 2024 | 06 | 5 | 65301 | 14 | 0 | 14 | 6 | 29 | 85 | 29 |
| 2024 | 07 | 5 | 82817 | 21 | 0 | 21 | 6 | 26 | 88 | 26 |
| 2024 | 08 | 5 | 135556 | 15 | 3 | 18 | 3 | 28 | 89 | 28 |
| 2024 | 09 | 4 | 63750 | 17 | 6 | 23 | 3 | 23 | 95 | 23 |
| 2024 | 10 | 5 | 43952 | 15 | 11 | 26 | 5 | 32 | 139 | 32 |
| 2024 | 11 | 5 | 64122 | 14 | 5 | 19 | 4 | 26 | 108 | 26 |

| | | | | | | | | | | |
|---|---|---|---|---|---|---|---|---|---|---|
| 2024 | 12 | 5 | 48948 | 7 | 12 | 19 | 4 | 23 | 142 | 23 |
| 2024 | 13 | 5 | 27055 | 11 | 9 | 20 | 2 | 40 | 141 | 40 |
| 2024 | 14 | 5 | 29586 | 14 | 5 | 19 | 5 | 28 | 126 | 28 |
| 2024 | 15 | 5 | 64617 | 9 | 6 | 15 | 0 | 20 | 123 | 20 |
| 2024 | 16 | 5 | 25589 | 10 | 0 | 10 | 2 | 17 | 107 | 17 |
| 2024 | 17 | 5 | 35975 | 6 | 0 | 6 | 2 | 22 | 77 | 22 |
| 2024 | 18 | 5 | 47400 | 6 | 0 | 6 | 3 | 23 | 51 | 23 |
| 2024 | 19 | 5 | 85515 | 4 | 0 | 4 | 3 | 31 | 79 | 31 |
| 2024 | 20 | 5 | 65672 | 7 | 0 | 7 | 1 | 25 | 61 | 25 |
| 2024 | 21 | 5 | 72239 | 6 | 1 | 7 | 3 | 22 | 62 | 22 |
| 2024 | 22 | 5 | 76353 | 12 | 0 | 12 | 2 | 17 | 52 | 17 |
| 2024 | 23 | 5 | 58478 | 9 | 0 | 9 | 5 | 27 | 75 | 27 |
| 2024 | 24 | 5 | 72081 | 13 | 0 | 13 | 6 | 28 | 64 | 28 |
| 2024 | 25 | 5 | 86478 | 11 | 0 | 11 | 4 | 18 | 67 | 18 |
| 2024 | 26 | 5 | 78474 | 9 | 0 | 9 | 2 | 22 | 54 | 22 |
| 2024 | 27 | 5 | 89174 | 9 | 0 | 9 | 3 | 28 | 53 | 28 |
| 2024 | 28 | 5 | 71811 | 8 | 1 | 9 | 3 | 19 | 48 | 19 |
| 2024 | 29 | 5 | 46024 | 22 | 5 | 27 | 3 | 32 | 100 | 32 |
| 2024 | 30 | 5 | 83677 | 12 | 1 | 13 | 3 | 28 | 80 | 28 |
| 2024 | 31 | 5 | 185984 | 18 | 0 | 18 | 6 | 28 | 82 | 28 |
| 2024 | 32 | 5 | 152959 | 19 | 1 | 20 | 9 | 25 | 56 | 25 |
| 2024 | 33 | 5 | 129177 | 21 | 6 | 27 | 8 | 24 | 119 | 24 |
| 2024 | 34 | 5 | 144531 | 28 | 1 | 29 | 8 | 14 | 60 | 14 |
| 2024 | 35 | 5 | 180309 | 17 | 5 | 22 | 7 | 31 | 87 | 31 |
| 2024 | 36 | 5 | 111348 | 12 | 2 | 14 | 5 | 25 | 85 | 25 |
| 2024 | 37 | 5 | 59773 | 13 | 1 | 14 | 8 | 24 | 62 | 24 |
| 2024 | 38 | 5 | 62511 | 13 | 0 | 13 | 2 | 15 | 45 | 15 |
| 2024 | 39 | 4 | 64145 | 12 | 0 | 12 | 4 | 32 | 70 | 32 |
| 2024 | 40 | 5 | 35878 | 7 | 0 | 7 | 2 | 18 | 59 | 18 |
| 2024 | 41 | 5 | 36485 | 6 | 0 | 6 | 2 | 16 | 58 | 16 |
| 2024 | 42 | 5 | 51753 | 5 | 1 | 6 | 1 | 26 | 66 | 26 |
| 2024 | 43 | 5 | 19803 | 4 | 1 | 5 | 1 | 26 | 67 | 26 |
| 2024 | 44 | 5 | 37693 | 9 | 0 | 9 | 3 | 16 | 57 | 16 |
| 2024 | 45 | 5 | 28092 | 5 | 0 | 5 | 1 | 24 | 58 | 24 |
| 2024 | 46 | 5 | 38892 | 2 | 0 | 2 | 1 | 19 | 50 | 19 |
| 2024 | 47 | 5 | 52460 | 8 | 0 | 8 | 4 | 16 | 55 | 16 |
| 2024 | 48 | 5 | 31489 | 7 | 2 | 9 | 4 | 19 | 72 | 19 |
| 2024 | 49 | 5 | 61032 | 10 | 6 | 16 | 4 | 29 | 125 | 29 |
| 2024 | 50 | 5 | 75193 | 13 | 9 | 22 | 4 | 29 | 124 | 29 |
| 2024 | 51 | 5 | 145218 | 25 | 7 | 32 | 9 | 53 | 213 | 53 |

| | | | | | | | | | | |
|---|---|---|---|---|---|---|---|---|---|---|
| 2024 | 52 | 5 | 206738 | 31 | 3 | 34 | 13 | 51 | 150 | 51 |
| 2025 | 02 | 5 | 185430 | 16 | 2 | 18 | 5 | 39 | 121 | 39 |
| 2025 | 03 | 5 | 196340 | 13 | 3 | 16 | 5 | 43 | 138 | 43 |
| 2025 | 04 | 5 | 134933 | 9 | 0 | 9 | 2 | 36 | 117 | 36 |
| 2025 | 05 | 5 | 165879 | 11 | 2 | 13 | 4 | 27 | 89 | 27 |
| 2025 | 06 | 5 | 43189 | 14 | 1 | 15 | 7 | 30 | 100 | 30 |
| 2025 | 07 | 5 | 90801 | 14 | 0 | 14 | 2 | 28 | 88 | 28 |
| 2025 | 08 | 5 | 69695 | 18 | 1 | 19 | 6 | 27 | 74 | 27 |
| 2025 | 09 | 5 | 65046 | 9 | 4 | 13 | 4 | 25 | 83 | 25 |
| 2025 | 10 | 5 | 59906 | 9 | 10 | 19 | 3 | 20 | 159 | 20 |
| 2025 | 11 | 5 | 70949 | 8 | 8 | 16 | 1 | 36 | 145 | 36 |
| 2025 | 12 | 5 | 43632 | 9 | 6 | 15 | 2 | 21 | 130 | 21 |
| 2025 | 13 | 5 | 46975 | 1 | 0 | 1 | 0 | 12 | 58 | 12 |

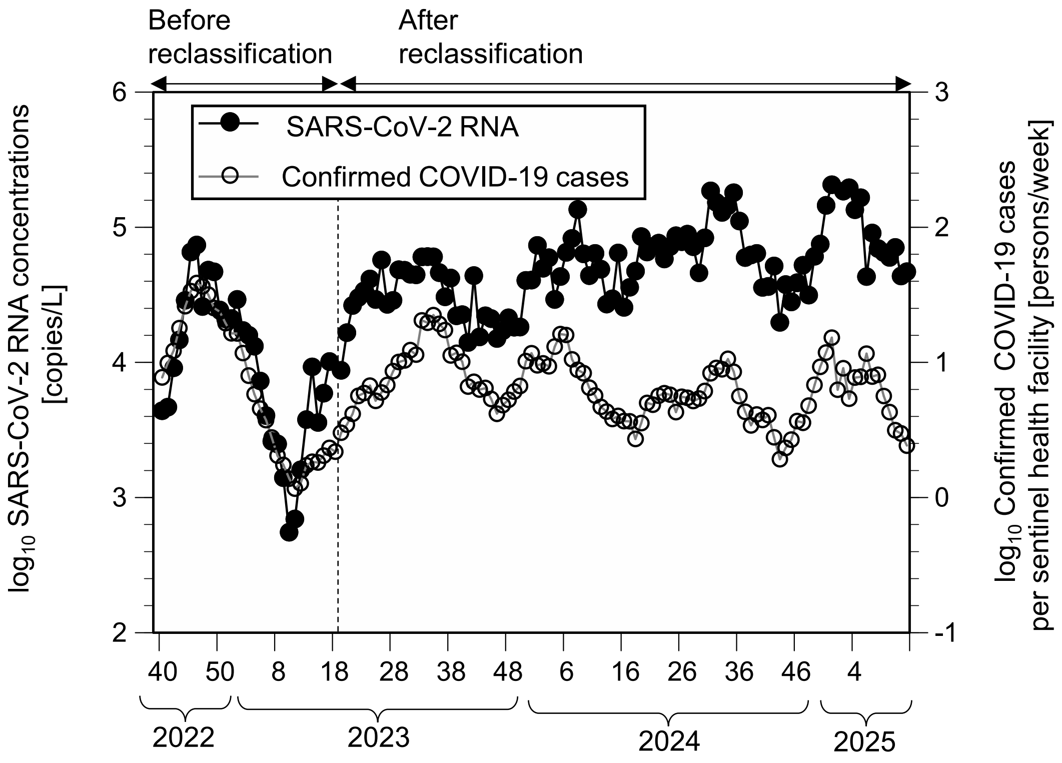


Figure S1. Temporal changes of SARS-CoV-2 concentrations in wastewater and confirmed COVID-19 cases per sentinel health facility in the City of Sapporo. Data sources were as follows: SARS-CoV-2 concentrations in wastewater during Weeks 9–17 of 2023 (Murakami et al., 2024); SARS-CoV-2 concentrations in wastewater in other weeks (present study); confirmed COVID-19 cases per sentinel health facility before reclassification (Murakami et al., 2024); confirmed COVID-19 cases per sentinel health facility after reclassification (City of Sapporo, 2025).


City of Sapporo, 2025. https://www.city.sapporo.jp/eiken/infect/trend/graph/l567.html (accessed August 8, 2025). (in Japanese)

Murakami, M., Ando, H., Yamaguchi, R., Kitajima, M., 2024. Evaluating survey techniques in wastewater-based epidemiology for accurate COVID-19 incidence estimation. Sci. Total Environ. 954, 176702.

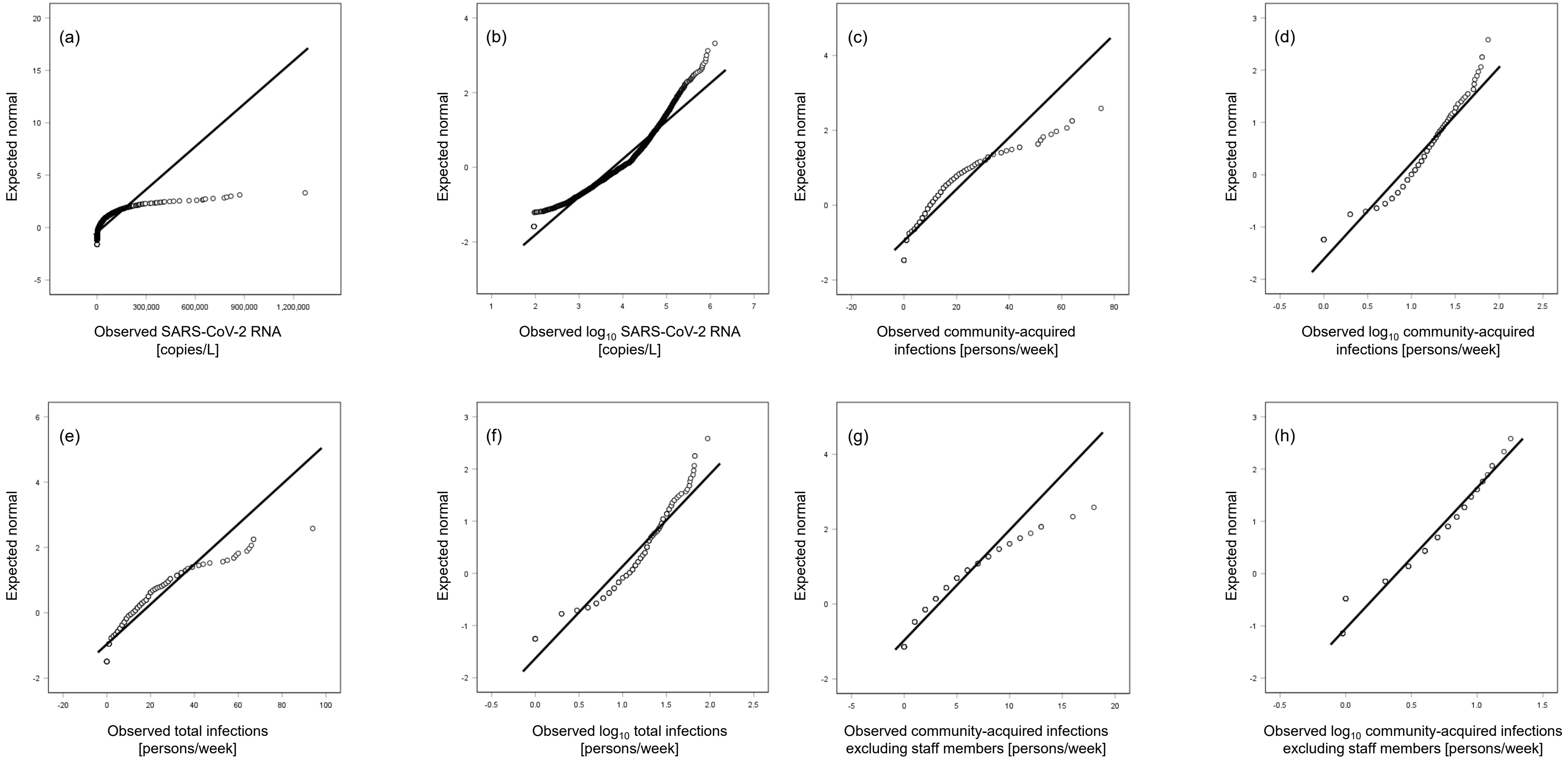


Figure S2. Q–Q plots for SARS-CoV-2 RNA concentrations, community-acquired infections, and total infections. (a) SARS-CoV-2 RNA concentrations, (b) $Log_{10}$ SARS-CoV-2 RNA concentrations, (c) Community-acquired infections, (d) $Log_{10}$ community-acquired infections, (e) Total infections, (f) $Log_{10}$ total infections, (g), Community-acquired infections, excluding staff members, (h) $Log_{10}$ community-acquired infections, excluding staff members.

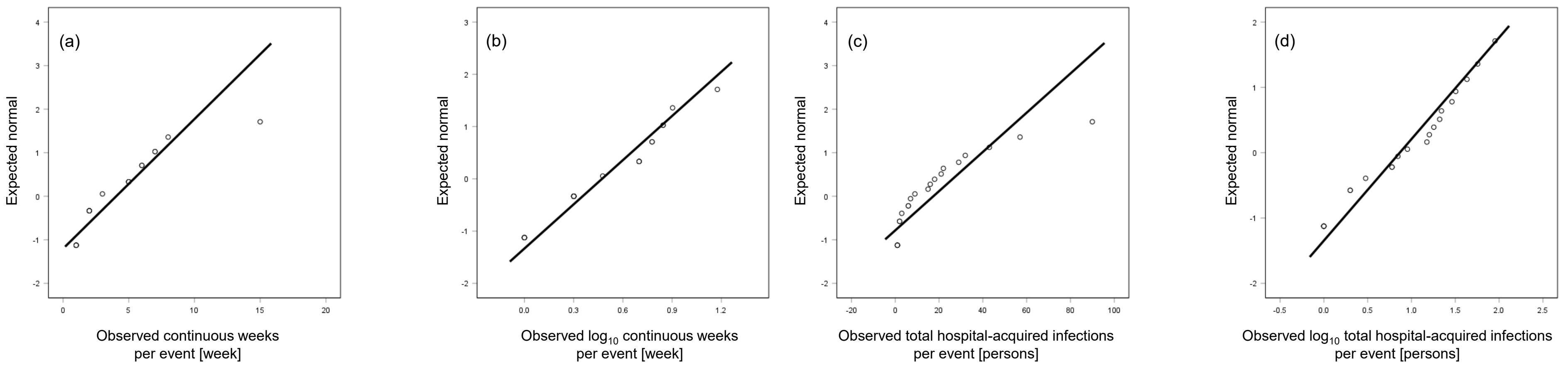


Figure S3. Q–Q plots for continuous week and total hospital-acquired patients per event. (a) Continuous week, (b) $\text{Log}_{10}$ continuous week, (c) Total hospital-acquired infections, (d) $\text{Log}_{10}$ total hospital-acquired infections.

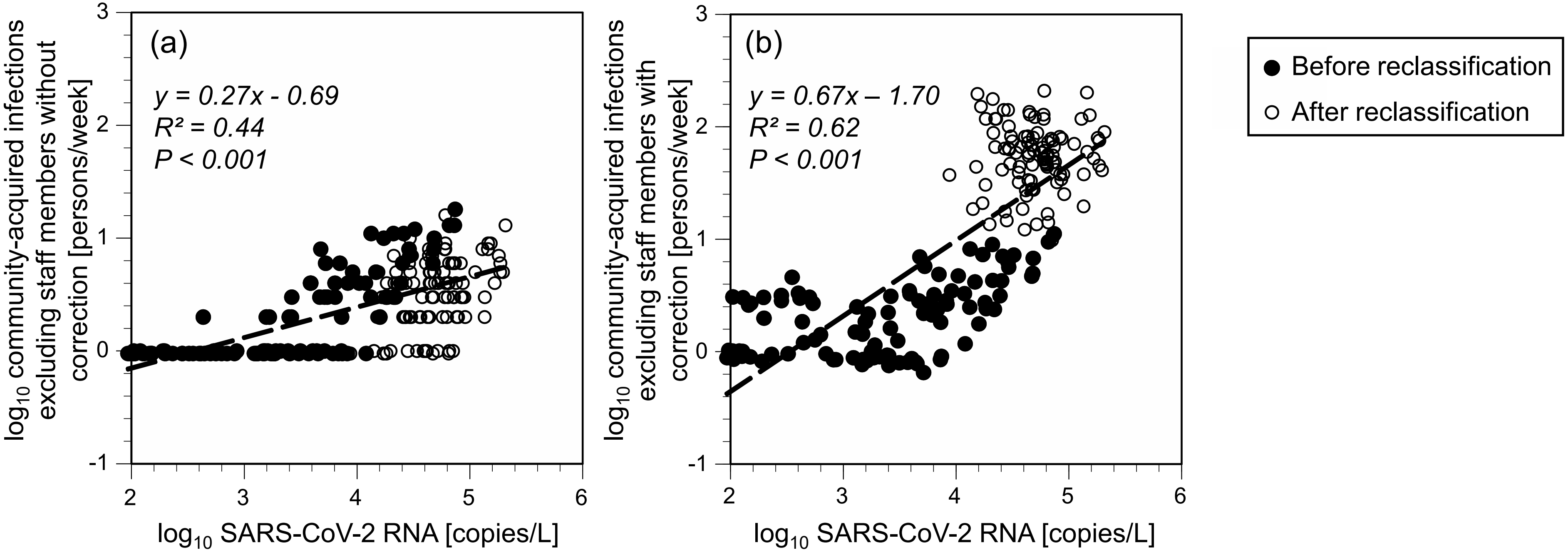


Figure S4. Scatterplots of SARS-CoV-2 RNA concentration vs. confirmed COVID-19 cases. (a) Community-acquired infections, excluding staff members without correction, by testing rate excluding staff members, (b) Community-acquired infections, excluding staff members, with correction, by testing rate excluding staff members.
